# Artificial intelligence applied on chest X-ray can aid in the diagnosis of COVID-19 infection: a first experience from Lombardy, Italy

**DOI:** 10.1101/2020.04.08.20040907

**Authors:** Isabella Castiglioni, Davide Ippolito, Matteo Interlenghi, Caterina Beatrice Monti, Christian Salvatore, Simone Schiaffino, Annalisa Polidori, Davide Gandola, Cristina Messa, Francesco Sardanelli

## Abstract

**Objectives:** We tested artificial intelligence (AI) to support the diagnosis of COVID-19 using chest X-ray (CXR). Diagnostic performance was computed for a system trained on CXRs of Italian subjects from two hospitals in Lombardy, Italy.

**Methods:** We used for training and internal testing an ensemble of ten convolutional neural networks (CNNs) with mainly bedside CXRs of 250 COVID-19 and 250 non-COVID-19 subjects from two hospitals. We then tested such system on bedside CXRs of an independent group of 110 patients (74 COVID-19, 36 non-COVID-19) from one of the two hospitals. A retrospective reading was performed by two radiologists in the absence of any clinical information, with the aim to differentiate COVID-19 from non-COVID-19 patients. Real-time polymerase chain reaction served as reference standard.

**Results:** At 10-fold cross-validation, our AI model classified COVID-19 and non COVID-19 patients with 0.78 sensitivity (95% confidence interval [CI] 0.74–0.81), 0.82 specificity (95% CI 0.78–0.85) and 0.89 area under the curve (AUC) (95% CI 0.86–0.91). For the independent dataset, AI showed 0.80 sensitivity (95% CI 0.72–0.86) (59/74), 0.81 specificity (29/36) (95% CI 0.73–0.87), and 0.81 AUC (95% CI 0.73– 0.87). Radiologists’ reading obtained 0.63 sensitivity (95% CI 0.52–0.74) and 0.78 specificity (95% CI 0.61–0.90) in one centre and 0.64 sensitivity (95% CI 0.52–0.74) and 0.86 specificity (95% CI 0.71–0.95) in the other.

**Conclusions:** This preliminary experience based on ten CNNs trained on a limited training dataset shows an interesting potential of AI for COVID-19 diagnosis. Such tool is in training with new CXRs to further increase its performance.

**Key points:**

- Artificial intelligence based on convolutional neural networks was preliminary applied to chest-X-rays of patients suspected to be infected by COVID-19.
- Convolutional neural networks trained on a limited dataset of 250 COVID-19 and 250 non-COVID-19 were tested on an independent dataset of 110 patients suspected for COVID-19 infection and provided a balanced performance with 0.80 sensitivity and 0.81 specificity.
- Training on larger multi-institutional datasets may allow this tool to increase its performance.

## Introduction

According to the John Hopkins Coronavirus Resource Centre [1], on April 3^rd^, 2020 the novel Coronavirus (COVID-19) infected more than 1,000,000 individuals with more than 50,000 deaths worldwide. The United States represent the most infected country, with a total of 259,750 cases and 6,603 deaths. Italy comes, being the most involved country in Europe, and the first country for number of deaths worldwide [2]. Other European countries are following the Italian trend with a delay of some weeks. In particular, since February 21th 2020, an increasing number of COVID-19 cases has been found in Lombardy, an Italian region where on April 2^nd^, 2020 data are 47,520 cases and 8,311 deaths [3].

In this pandemic situation, clinicians are dramatically requesting fast diagnostic tools for COVID-19 characterized by a good balance between sensitivity and specificity, leading to acceptable predictive values in a context of a variable prevalence, depending on policies ranging from testing only symptomatic subjects to mass screening. Of note, any tool to be applied for this aim should have a good cost-benefit ratio for the healthcare service.

The reference standard for COVID-19 is reverse transcriptase polymerase chain reaction (RT-PCR) [4], even though this test can give a false negative result at an early stage of the disease and the time needed to get its result is highly variable. At any rate, considering the most relevant clinical evolution leading to pneumonia, chest imaging study are routinely performed in suspected or confirmed COVID-19 cases [5].

Computed tomography (CT) studies showed that the most common imaging finding in COVID-19 are ground-glass opacities (GGOs) scattered throughout the lungs. This finding is given by air-sacs or alveoli filled with fluid, represented as a shade of grey on images [6]. In more advanced disease, GGOs progress to “consolidations”. Swelling of the interstitial space along the walls of the lung lobules can create the “crazy paving” aspect: walls appear thicker as white lines over the GGO background. These three findings (GGOs, consolidations, and crazy paving) can be isolated or combined, with GGOs being commonly the first sign.

In COVID-19 patients these findings usually involve multiple lobes bilaterally, frequently affecting the peripheral posterior lungs but in early phases of the disease, one lobe can be affected [4].

A large study on 1,014 Chinese patients [7] showed that while the sensitivity of initial RT-PCR was only 59%, that of initial chest CT was 97%. However, CT specificity was only 25%, while the positive predictive value was 65% and the negative predictive value 83%. In addition, we should consider that the availability of CT as first performed imaging test for all suspected COVID-19 patients is not easy to implement, also considering the time needed for sanitizing the room and the CT equipment after examination [8].

When a patient has symptoms of COVID-19, like fever, cough, or dyspnoea, chest X-ray (CXR) is usually the first imaging performed, because it is cheaper and easier to do. Furthermore, CXR can also be acquired with portable instrumentation in isolated rooms in emergency departments or at the patient’s bedside in every other department, which would considerably ease the required sanitization process. CXR images have a high spatial resolution, but they are planar images, not allowing three-dimensional slicing as. Anyway, even for CXR the most common reported abnormal finding are GGOs, with portions of the lungs appearing as a “hazy” shades of grey instead of being black with fine white lung markings for blood vessels [9]. As GGOs are usually the first radiological sign of COVID-19 pneumonia, it could be hypothesized to be able to improve the early diagnosis of COVID-19 by means of a smarter reading of CXRs.

Artificial intelligence (AI) is emerging as unique powerful method to improve diagnosis and prognosis of several multifactorial diseases, including pneumonia. In 2018, a worldwide competition on the Kaggle portal (www.kaggle.com) was launched by the Radiological Society of North America on the complex task of automatically screening pneumonia (viral and bacterial) [10] versus non-pneumonia patients on CXRs. Many research groups participated and trained their AI algorithms on thousands of CXRs images with clinical diagnoses of viral or bacterial pneumonia and non-pneumonia made available from several US hospitals. Leader groups obtained excellent results training their different AI systems on CXRs [11]. More recently, a Chinese research team proved the potential of AI in supporting the diagnosis of COVID-19 in Chinese population suspected by COVID-19 when trained on CT images, showing excellent results, with sensitivity and specificity higher than 90% [12]. However, their AI CT-based model may not be implemented in an emergency context as for COVID-19 pandemic.

Thus, the aim of our study was to test AI applied to CXRs in the COVID-19 emergency setting also considering the radiologists’ reading performance. Our purpose was to develop a tool able to support the diagnosis of COVID-19, offering a second opinion to clinical radiologists worldwide.

## Methods

### Patient population

Approval of the study has been obtained from the local Ethics Committee (Ethics Committee of IRCCS San Raffaele). In our retrospective study we used a case-control design based on non-consecutive patients and an artificially enriched positive class (COVID-19).

### Training and internal testing set

We included in the training set the CXRs of the following groups. First, non-consecutive patients suspected of COVID-19 infection admitted to the Hospital San Gerardo, Monza, Italy (centre 1) from 1^st^ March to 13^th^ March 2020 (n = 270, 135 COVID-19 and 135 non-COVID-19.). Thus, we artificially enriched the positive class (COVID-19) of the case-control study for balanced training.

Clinical suspicion of COVID-19 was defined upon arrival at the emergency room and based on referring physician’s judgment for patients admitted at the emergency department, taking into consideration: onset of symptoms (the main fever, cough and dyspnoea) and blood tests (white blood cell count, red blood cell count, C-reactive protein level). The most common symptoms were fever (87.2%), followed by cough (56.2%) and dyspnoea (40.3%). These patients underwent digital CXR in anteroposterior projection at bedside as well as real-time RT-PCR assays using commercial kits (ribonucleic acid was extracted from collected samples). The classification of positive or negative COVID-19 cases was based on the detection or non-detection of the pathogen: the number of cases of negative RT-PCR followed by one or more further swabs was 135.

Second, we included in the training dataset digital CXRs of consecutive patients suspected for COVID-19 infection according to the same criteria, admitted to the IRCCS Policlinico San Donato (centre 2) from February 25^th^ to March 16^th^ 2020 and subsequently confirmed to be COVID-19 infected by RT-PCR (n = 115), thus composing an artificially enriched positive class for balanced training. Out of these CXRs, 87 were anteroposterior projections performed at bedside and 28 posteroanterior projections acquired in upright position. Third, the dataset was enriched with CXRs of non-consecutive sex- and age-matched patients admitted to IRCCS Policlinico San Donato approximately in the same time interval of the previous year (n = 115, February 15^th^ to March 16^th^ 2019) without any mention of lung abnormalities in the radiological report, 16 of them being anteroposterior projections performed at bedside, 99 of them being posteroanterior projections acquired in upright position.

### External testing set

We then retrospectively considered consecutive patients suspected of COVID-19 infection admitted to the Hospital San Gerardo, Monza (MI) (centre 1) from 14^th^ March to 19^th^ March 2020, thus temporally separated from the training set coming from the same centre. Clinical suspicion of COVID-19, digital bedside CXRs and specific RT-PCR assays were performed as previously described (n = 110), 74 of them resulted to be COVID-19 infected and 36 non-confirmed at RT-PCR assay.

A flow diagram describing patient selection is depicted in Fig. 1, while Table 1 summarizes included patients’ provenience and COVID-19 positivity or negativity.

**Table 1:**
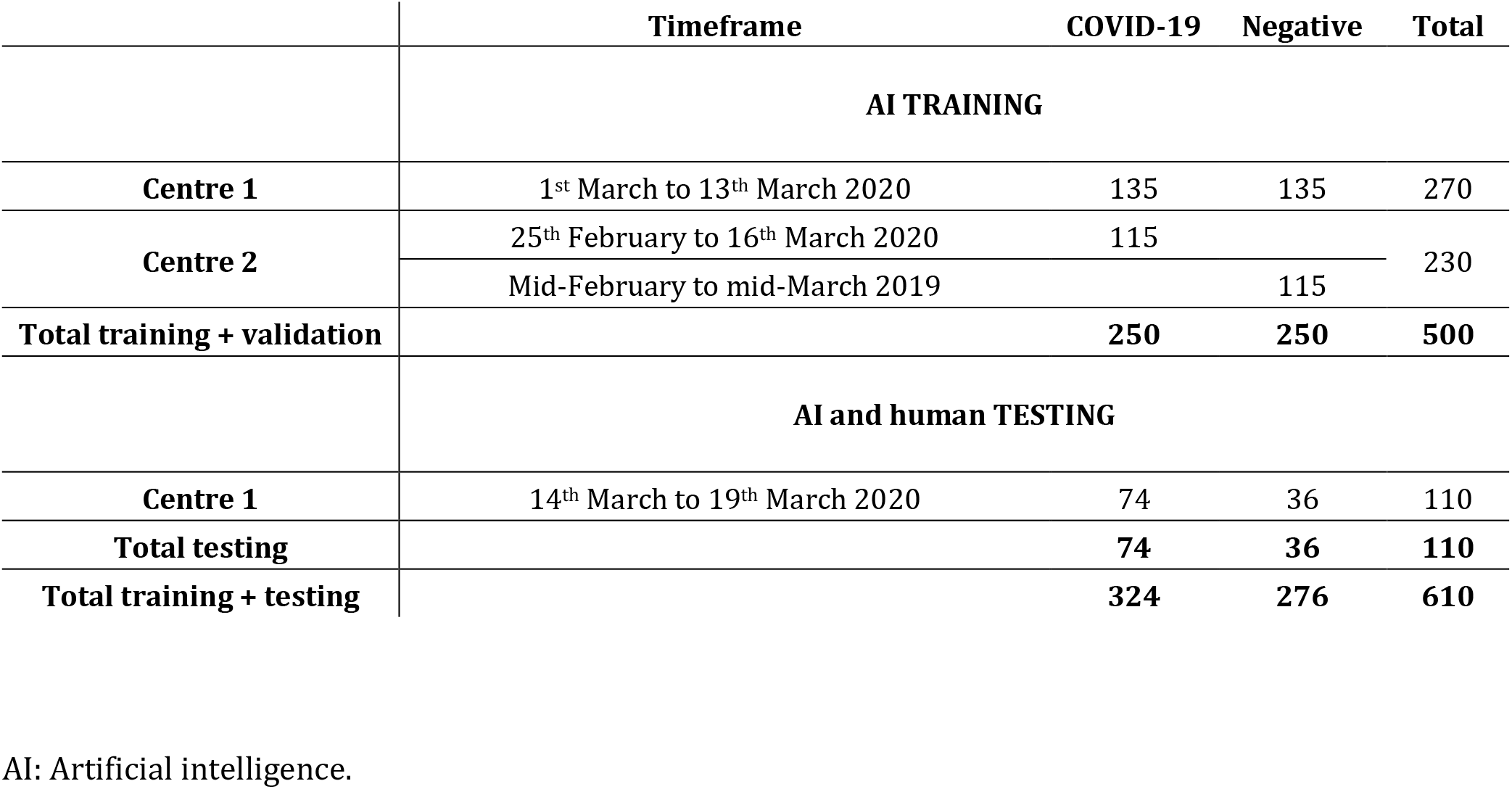
Provenience and characteristics of the included chest X-ray exams.

|  | Timeframe | COVID-19 | Negative | Total |
| --- | --- | --- | --- | --- |
| **AI TRAINING** |  |  |  |  |
| **Centre 1** | 1<sup>st</sup> March to 13<sup>th</sup> March 2020 | 135 | 135 | 270 |
| **Centre 2** | 25<sup>th</sup> February to 16<sup>th</sup> March 2020 | 115 |  | 230 |
|  | Mid-February to mid-March 2019 |  | 115 |  |
| **Total training + validation** |  | **250** | **250** | **500** |
| **AI and human TESTING** |  |  |  |  |
| **Centre 1** | 14<sup>th</sup> March to 19<sup>th</sup> March 2020 | 74 | 36 | 110 |
| **Total testing** |  | **74** | **36** | **110** |
| **Total training + testing** |  | **324** | **276** | **610** |

AI: Artificial intelligence.

**Figure 1:**
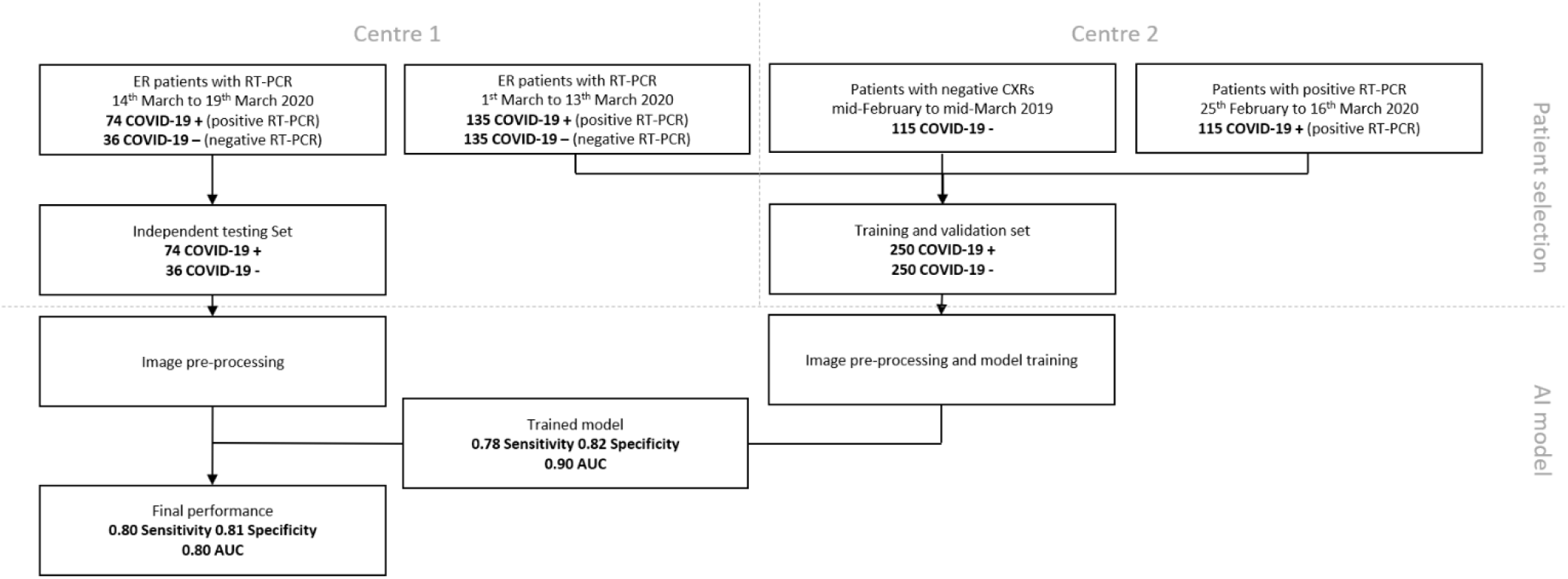
**Fig. 1** Flow diagram for patient selection. CXR: chest X-ray; RT-PCR: reverse transcriptase polymerase chain reaction; ER: emergency room; AUC: area under the curve. CXR: chest X-ray; RT-PCR: reverse transcriptase polymerase chain reaction; ER: emergency room; AUC: area under the curve.

### Image analysis by AI

For this purpose, the TRACE4© radiomic platform was used (http://www.deeptracetech.com/tempo/TechnicalSheetTRACE4.pdf) to train, validate and test different AI systems combined with different feature extraction methods. The TRACE4© platform includes: a) full workflow for radiomic analysis (compliant to the guidelines of International Biomarker Standardization Initiative, IBSI), b) different feature extraction and selection methods, and c) different ensembles of machine-learning techniques such as support vector machines, random forests, deep learning and transfer learning of neural networks. All these functions are fully integrated in the TRACE4© software allowing an easy development and implementation of AI models based on medical images also to non-expert users. In addition, TRACE4© includes ready-to-used AI models for classification of different complex diseases (e.g. Alzheimer’s diseases, breast cancer, ovarian cancer).

Starting from the included CXR images, we trained and tested an ensemble of 10 convolutional neural networks (ResNET50) [13] with sum vote rule (ensemble-averaging of class probability) using a 10-fold cross-validation method. The classification task of interest was binary (COVID-19 versus non COVID-19), considering COVID-19 all patients with positive RT-PCR (250) from both centres and non-COVID-19 all patients with negative RT-PCR from centre 1 and those admitted to centre 2 for CXR in the same period the previous year before with a CXR reported as negative (see Table 1).

The proposed deep-neural-network classifier adopted the ResNet-50 architecture, a convolutional neural network composed of 50 layers, each coupled with two convolutional filters that is able to learn a rich feature representation of the input classes (more than a million of images from the ImageNet database) (ImageNet. http://www.image-net.org), and to use this feature representation to classify new images as belonging to one of the input classes layers, and a fine-tuning process was applied to the original ResNet-50 architecture to specialize its last layers to the binary discrimination task (COVID-19 vs non COVID-19).

CXR images were fed into the deep neural network with a pre-processing down-sampling of image size of about 1/5 of the original image size (depending from the different original image size). In order to increase X-ray image diversity among different training phases (epochs), data-augmentation techniques (including image manipulations such as rotation and cropping) were applied to the CXR set during the training of the classifier. The maximum number of epochs was set to 30, with a mini-batch size of 8 (the samples of the training set were randomized before each epoch in order to avoid issues related to the choice of samples to include in the mini-batches - e.g. always discarding the same samples). A schematic drawing for the network architecture is presented in Fig. 2.

**Figure 2:**
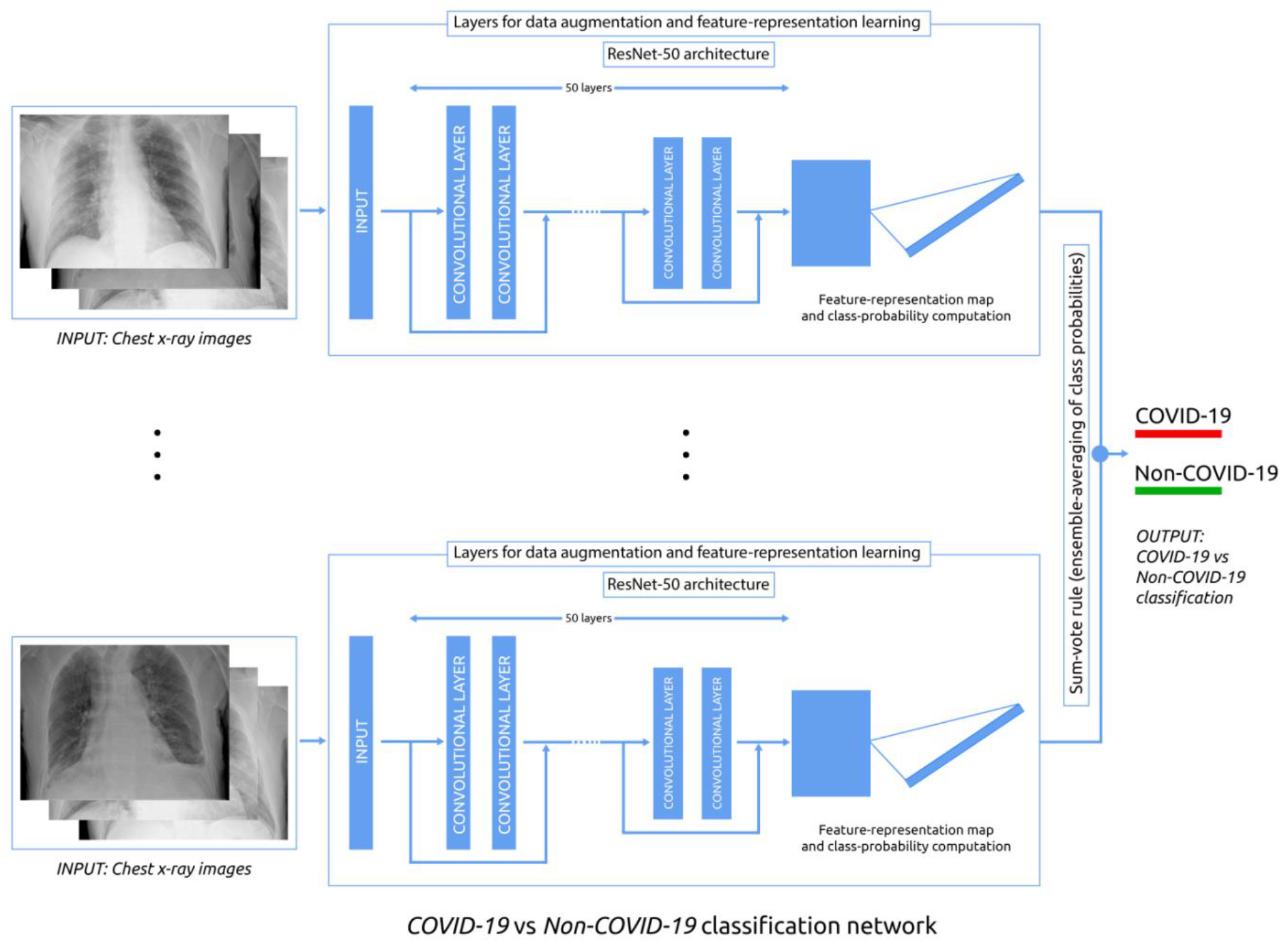
Schematic drawing of the AI-network architecture for the classification of Covid-19 vs Non-Covid-19 patients through CXR imaging.

The performance obtained by our AI system are presented in terms of: accuracy for training, sensitivity, specificity, area under the curve (AUC), positive predictive value (PPV), negative predictive value (NPV) for both cross validation and independent testing (for training and cross-validation, with standard deviation and corresponding 95% confidence interval [CI]).

### Image analysis by radiologists

A retrospective reading of CXRs was performed by staff radiologists at both hospitals. They were one radiologist with 15 years of experience in chest imaging at centre 1 (reader 1) and a general radiologist with 6 years of experience at the centre 2 (reader 2). They were asked to standardize the reading without any information on medical history, clinical and biologic data, with aim to differentiate COVID-19 patients from non-COVID-19 patients. Both the readers assessed the external testing cases consisting into 110 bedside CXRs of patients suspected to be COVID-19 infected, all from the emergency department of centre 1 from March 16 to March 19 2020, 74 of them finally resulting positive for COVID-19 at RT-PCR and 36 negative for COVID-19 at RT-PCR (see Table 1).

Results of human reading performance (sensitivity, specificity, PPV and NPV) were computed and presented as ratios with their 95% CI.

## Results

### Image analysis by AI

Accuracy for the training was 0.99 ± 0.01 (95% CI 0.98-1.00). For internal testing (cross validation) of CXRs (250 of COVID-19 and 250 of non-COVID-19 subjects), our AI model was able to automatically classify patients with sensitivity of 0.78 ± 0.09 (95% CI 0.74–0.81), specificity of 0.82 ± 0.11 (95% CI 0.78– 0.85), PPV of 0.81 ± 0.08 (95% CI 0.78–0.85), NPV of 0.79 ± 0.08 (95% CI 0.76–0.83), and AUC of 0.89 ± 0.04 (95% CI 0.86-0.91), respectively (10-fold cross validation). For the 110 CXRs of the independent (temporally separated) group of suspect COVID-19 patients, our AI system showed sensitivity of 0.80 (95% CI 0.72–0.86), specificity of 0.81 (95% CI 0.73–0.87), PPV of 0.89 (95% CI 0.82–0.94), NPV of 0.66 (95% CI 0.57–0.75), and AUC of 0.81 (95% CI 0.73–0.87). To be noted, PPV and NPV of the independent test were affected by the imbalance classes (while this did not occur in internal testing). Table 2 shows a comprehensive list of the performance obtained by the AI model.

**Table 2.** Results for the artificial intelligence and human readings of study datasets. Diagnostic performance data are presented as value and 95% confidence interval. AI: artificial intelligence; PPV: positive predictive value; NPV: negative predictive value.

| Covid-19 vs Negative<br>AI validation (cross-validation) |  |  |
| --- | --- | --- |
|  | Positive (n) | Negative (n) |
| Assigned positive | 195 | 46 |
| Assigned negative | 55 | 204 |
|  | **Sensitivity**<br>0.78** (0.74-0.81) | **Specificity**<br>0.82** (0.78-0.85) |
|  | **PPV**<br>0.82** (0.78-0.85) | **NPV**<br>0.80** (0.76-0.83) |
| AI independent testing |  |  |
|  | Positive (n) | Negative (n) |
| Assigned positive | 59 | 7 |
| Assigned negative | 15 | 29 |
|  | **Sensitivity**<br>0.80** (0.72-0.86) | **Specificity**<br>0.81** (0.73-0.87) |
|  | **PPV** | **NPV** |
|  | 0.89** (0.82-0.94) | 0.66** (0.57-0.75) |
| **Human independent testing (reader 1)** |  |  |
|  | **Positive (n)** | **Negative (n)** |
| **Assigned positive** | 47 | 8 |
| **Assigned negative** | 27 | 28 |
|  | **Sensitivity** | **Specificity** |
|  | 0.64 (0.52-0.74) | 0.78 (0.61-0.90) |
|  | **PPV** | **NPV** |
|  | 0.85 (0.76-0.92) | 0.51 (0.42-0.59) |
| **Human independent testing (reader 2)** |  |  |
| **Assigned positive** | 47 | 5 |
| **Assigned negative** | 27 | 31 |
|  | **Sensitivity** | **Specificity** |
|  | 0.64 (0.52-0.74) | 0.86 (0.71-0.95) |
|  | **PPV** | **NPV** |
|  | 0.90 (0.80-0.96) | 0.53 (0.61-0.79) |

[\* = p-value < 0.05; \*\* = p-value < 0.005]

### Image analysis by radiologists

For the 110 cases from centre 1 used for the external testing of the AI system, human reader 1 showed sensitivity of 0.64 (95% CI 0.52–0.74), specificity of 0.78 (95% CI 0.61–0.90), PPV of 0.85 (95% CI 0.76– 0.93) and NPV of 0.51 (95% CI 0.42–0.59); human reader 2 showed sensitivity of 0.64 (95% CI 0.52–0.74), specificity of 0.86 (95% CI 0.71–0.95), PPV of 0.90 (95% CI 0.80–0.96), NPV of 0.53 (0.61–0.79) (see Table 2).

## Discussion

Novel coronavirus pneumonia (COVID-19) is a viral infectious disease transmitted through air droplets and close distance contacts. The outbreak of COVID-19 epidemic has resulted in a global health emergency, more diffuse than the coronavirus severe acute respiratory syndrome (SARS) in 2003, both caused by viruses belonging to the Coronaviridae family (3). As a matter of fact, on March 13, 2020, the WHO declared the COVID-19 outbreak a pandemic [14].

Diagnosing the disease quickly and accurately is a clinical need and CXR is a vital diagnostic tool for COVID-19 associated pneumonia in emergency. However, its performance in the diagnosis of COVID-19 cases has not yet been reported by large studies. This study collected a total of 495 COVID-19 patients who had CXR with a positive RT-PCR, enriched with other 115 non-COVID-19 patients with CXR in an equivalent period preceding the epidemic, in order to train and test a CNN-based AI system.

The main finding of our study is that the performance of our AI system, even though trained on a limited number of cases (500, 250 of them COVID-19 positive and 250 COVID-19 negative), showed an interesting performance: at 10-fold cross-validation, the sensitivity was 0.78 and the specificity was 0.82. When challenged on an independent new dataset (110 cases, 74 of them COVID-19 positive and 36 COVID-19 negative), the system showed a sensitivity of 0.80 and a specificity 0.81. To obtain a reference estimate, we asked human readers to assess these 110 CXRs aiming at differentiating COVID-19 from non-COVID-19 patients in suspected to be infected by this virus, being blinded to any medical history as well as clinical and biological data. The two radiologists, coming from two different hospitals, obtained exactly the same limited sensitivity (0.64) and similar specificity, as shown by the large overlapping of the 95%CIs.

It is highly likely that a human reading completely informed about history and clinical data or during booming of the epidemic with an increasing prevalence would have been able to strongly increase the sensitivity, but a trade-off could be paid in terms of specificity. This phenomenon is well visible in the case of CT in the recent report by Ai et al [7] where a 97% sensitivity is counterbalanced by a 25% specificity.

The performance of our AI system appears interesting for the well balance between the two terms, with 0.80 sensitivity and 0.81 specificity.

This constitutes a promising starting point, especially when considering the technical issue regarding bedside CXRs that were evaluated by the AI system: only one anteroposterior projection in supine position. This means that there is room for improving CXR performance in these patients. On the one side, the AI system can be trained on thousands of cases, applying the deep-learning general principle: more data you use for training, a higher performance you get [15]. On the other side, CXR using the standard approach. i.e. both the posteroanterior and lateral projections to the patient standing in upright position, could substantially increase the quality of the radiograms and the three-dimensional information provided. However, this “state-of-the-art” approach is not always easy to carry on in the epidemic context, taking into consideration the possible contemporary use.

Since decision-making regarding 2019-COVID-19 management, especially diagnostic and treatment issues (home isolation or hospital admission, etc.), rely on the initial patient assessment, and because suspected COVID-2019 cases often are seen in the emergency department, developing strategies that improve early management is essential. To provide an improved diagnostic performance of CXR should be considered a goal, in particular when considering the difficulties of a CT strategy for early diagnosis, especially when the radiologist has not yet experienced in the reading of many cases, and in particular but not only in low-income regions.

Lastly, it is important to recognize that the role of CXR in patients’ evaluation depends on the severity of infection in the individual patient, as well as on the COVID-19 prevalence in the community. In individual who are asymptomatic or have mild disease, the sensitivity of CXR could fail if performed in the first 48 hours from the onset of symptoms. Individuals with very mild disease may eventually have positive RT-PCR results but would have been missed by early CXR. Conversely, CXR should be most useful in patients who are acutely ill and symptomatic in areas with relatively high prevalence, such as currently in Lombardy, in Italy. In this scenario, patients with clinical condition and CXR findings attributable to COVID-19 could be considered as possibly infected by the virus when the first RT-PCR test result is still not available or negative.

This study has some limitations. First, we trained our model on a limited number of cases, from the same geographical area. We could improve performances and generalizability of our model by adding new images, in particular from different geographical regions than Lombardy. Second, the external validation set was only temporally separate but not geographically separate from the training one and also relatively small. Third, we did not include other data such as clinical conditions such as symptoms and pulse oximeter data as complementary information to be given to the AI model and the human readers, a perspective to be explored in future studies.

In conclusion, we preliminarily showed that a CNN-based AI system applied to bedside CXR in patients suspected to be infected with COVID-19, even though trained on a limited number of cases, enable to reach a 0.80 sensitivity and a 0.81 specificity in an independent temporally separate patient group. The system could be used as a second opinion tool in studies aimed at assessing its usefulness for improving the final sensitivity and specificity in different geographical and temporal setting. Its performance could be improved by training on larger multi-institutional datasets.

## Data Availability

Data used in this work are not publicly available.

## Abbreviations

RT-PCR: Polymerase chain reaction
CT: Computed tomography
GGOs: Ground-glass opacities
CXR: Chest X-ray
AI: Artificial intelligence
AUC: Area under the curve
CNN: Convolutional neural network

## Notes

### Competing Interest Statement

F. Sardanelli has received research grants from and is member of speakers’ bureau and of
advisory group for General Electric, Bayer, and Bracco. Isabella Castiglioni, Matteo Interlenghi
and Annalisa Polidori own DeepTrace Technologies S.R.L shares. Christian Salvatore is CEO
of DeepTrace Technologies S.R.L. DeepTrace Technologies S.R.L is a spin-off of Scuola
Universitaria Superiore IUSS, Pavia, Italy.
The other authors of this manuscript declare no relationships with any companies, whose
products or services may be related to the subject matter of the article.

### Funding Statement

This research did not receive any specific grant from funding agencies in the public,
commercial, or not-for-profit sectors. This study was partially supported by funding from the
Italian Ministry of Health to IRCCS Ospedale San Raffaele.

